# Effects of a clinic-based reproductive empowerment intervention on reproductive coercion, intimate partner violence, and pregnancy outcomes: A cluster-controlled trial in Nairobi Kenya

**DOI:** 10.64898/2026.10.21.26366167

**Authors:** Jay G. Silverman, Jasmine Uysal, Chi-Chi Undie, Wilson Liambila, Lotus McDougal, Erin Pearson, Sabrina Boyce, Nicole Johns, Seri Wendoh

## Abstract

**Background:** To address reproductive coercion and intimate partner violence, the ARCHES (Addressing Reproductive Coercion in Health Settings) intervention was adapted for clinics in Nairobi, Kenya and evaluated to assess impacts on reproductive coercion (primary outcome), physical and sexual intimate partner violence (primary outcomes) and incident pregnancy (secondary outcome).

**Methods:** A parallel, non-blinded, non-randomized, cluster-controlled trial in six health clinics (1:1 allocation) was conducted. Clinics were allocated to receive ARCHES provider training or no additional training based on feasibility. Non-pregnant, non-sterilized female clients seeking family planning services were recruited. Inverse probability by treatment weighting (IPTW) used in difference-in-differences modified intent-to-treat (mITT) and as-treated approaches to assess the change in primary outcomes.

**Results:** A total of 659 women were recruited (328 ARCHES, 331 control). ARCHES was associated with significantly greater reductions in physical intimate partner violence compared to controls [mITT beta -0.98 (-1.58, -0.39), as-treated beta -0.56 (-1.19. 0.07)]. Both groups experienced reductions in reproductive coercion and sexual intimate partner violence decreased in both groups but controls showed greater reductions in reproductive coercion, and differences in sexual violence were not significant in as-treated models. ARCHES participants had lower odds of incident pregnancy at six-month follow-up [mITT AOR 0.42 (0.27, 0.67), as-treated AOR 0.29 (0.08. 1.02)].

**Conclusions:** ARCHES may improve women’s safety and reproductive agency in Nairobi. Further study is required to clarify effects on reproductive coercion and sexual intimate partner violence.

**Trial registration:** ClinicalTrials.gov [NCT03534401]

## Introduction

Intimate partner violence (IPV) is a global driver of unmet need for contraception and unintended pregnancy, impacting nearly one in three women around the world, particularly in low-middle income countries (LMICs).(1–9) Reproductive coercion (RC) is a form of gender-based violence comprised of behaviors by male partners or family members that create barriers to female access to and use of contraceptive methods and pregnancy decisions.(10) Though RC often co-occurs with IPV, it is a distinct form of violence that presents differently and can often be overlooked during IPV screening in the absence of a targeted approach. RC has been shown to be associated with poor reproductive health outcomes and unintended pregnancy, with some studies indicating these associations to be independent of IPV.(10, 11) Common forms of RC include pregnancy coercion (e.g., behaviors that force a woman to undergo a pregnancy she did not want), and contraceptive sabotage (e.g., tampering with a woman’s family planning method to reduce its effectiveness).(12, 13). While many forms of RC involve pregnancy promotion, it is important to note that such gender-based coercion may also take the form of pregnancy termination. For example, coercion of women by male partners and other family members to seek abortions against their will has been documented in multiple national contexts(14).

Kenya is an important setting in which to examine these gender inequities, having substantial gender equity and pregnancy prevention gaps, indicated by the high prevalence of IPV (30% physical, 10% sexual) and that more than two in five (43%) Kenyan women report their most recent pregnancy as unintended according to the 2022 Kenya DHS (15).

Governments and multilateral organizations have made broad calls to address gender-based violence, including RC and IPV, in routine sexual and reproductive health services,(1) with women in these settings more likely to have experienced IPV than those in the general population.(16) The Addressing Reproductive Coercion in Health Settings (ARCHES) program addresses this need by integrating strategies to address RC and IPV into routine reproductive health services. Core components of this single-session approach include education and counseling regarding RC (including strategies to regain contraception and pregnancy autonomy), opportunities to disclose RC and IPV, an educational palm-sized booklet with information on RC and IPV and local IPV services, and clinician-assisted referrals to local IPV services.(17) ARCHES has been demonstrated to reduce RC and IPV in the United States (18–20). In low- and middle-income countries (LMICs), ARCHES was first adapted to the Kenyan context (17). Modified versions of this original ARCHES LMIC adaptation(17) have been evaluated in Nigeria among women seeking either contraceptive or antenatal care and in Bangladesh among women seeking abortion care, with findings indicating that ARCHES reduces IPV in both contexts but reduces incident pregnancy only in Bangladesh and RC only in the Nigeria context(21, 22). Although both rigorous trials contribute critical evidence regarding impact of this approach, neither clarifies the impact of ARCHES on the population for which it was designed - women seeking routine contraceptive counseling. The current study of the original adaptation in Kenya also provides evidence of the impact of ARCHES in the East African context. As this model provides the basis for all subsequent ARCHES implementation in LMICs, this study also represents a critical reference point for this growing body of evidence.

The specific objectives of the current study are to assess the effects of the ARCHES intervention on the primary outcomes of RC and physical and sexual IPV and on the secondary outcomes of leaving an abusive relationship and incident and unintended pregnancy among women seeking contraceptive counseling in six community-based clinics in Nairobi, Kenya via a cluster-controlled trial.(17) Analysis of all outcomes is at the individual level. A cluster design was employed as providers are trained to modify their standard-of-care for all female patients with whom they are able to obtain privacy. Effects on contraceptive use (including covert use) and use of IPV services are reported elsewhere.(23)

## Materials and Methods

### Study Design

We conducted a six-month, prospective two-arm, parallel-group, non-blinded, non-randomized, cluster-control trial of the ARCHES intervention in six non-governmental organization (NGO) community-based clinics within metropolitan Nairobi, Kenya. The clinics were operated by Family Health Options Kenya (FHOK). All six clinics operated by FHOK in metropolitan Nairobi were selected for inclusion. No clinics were excluded. Intervention assignment was at the clinic level, as ARCHES provider training is expected to modify provider counseling and interactions with all patients. Clinics were assigned to intervention or control in a one-to-one allocation ratio based on feasibility of implementation (i.e., pilot clinics included as intervention) by the study team. FHOK approved inclusion of all clinic clusters. Matched pairs were attempted using available facility data on patient volume, patient socio-demographics, services provided and slum vs. non-slum location in an attempt to balance treatment groups; however, paired clinics were highly dissimilar due to outdated and inaccurate facility data and were, thus, disregarded for analysis. Providers at intervention clinic received a three-day training on the ARCHES intervention and implemented ARCHES within their existing model of contraceptive counseling, while control clinic providers received no additional training and continued to offer the same model of care. Data collection included surveys at baseline (prior to receiving service), exit (immediately after receiving service) and three- and six-month follow-up. Baseline and exit data were collected between July 30^th^ through December 15^th^, 2018; three-month follow-up data were collected from October 30, 2018, through April 1, 2019; and six-month follow-up data were collected from February 22^nd^ through June 30^th^, 2019. This trial was registered in ClinicalTrials.gov (ID: NCT03534401) on May 23, 2018.

### Participants

Participants were recruited at each of the six participating clinics and self-selected into intervention or control conditions based on the clinic at which they sought treatment in a convenience sample. When female patients checked into participating clinics, they were offered the opportunity to participate in a “women’s health study”. Women who expressed interest were then escorted by a trained research assistant (RA) to a private location within the clinic, unaccompanied by any other individuals, where they received a study overview and were screened for eligibility. Eligibility criteria consisted of having come to the clinic “for family planning or are interested in receiving family planning” on that day, self-reported being biologically female, between ages 15-49, not currently pregnant, not sterilized, having a male partner with whom they had sex in the past three months, able to safely participate in a private interview, planning to remain in the study geographic area for the next six months, and having a mobile phone safe for use to re-contact for a follow-up survey. Participants were excluded if they did not reporting having come to the clinic for family planning services, did not complete the exit survey immediately after their appointment before leaving the clinic, or reported on the exit survey that they did not receive family planning counseling at their appointment that day.

Eligible participants who agreed to participate provided written informed consent and completed interviewer-administered surveys by trained RAs using tablet computers in English or Kiswahili based on participant preference. Baseline surveys were conducted prior to contraceptive counseling, and exit surveys were conducted subsequent to contraceptive counseling but prior to participants’ leaving the clinic. Participants were re-contacted for three and six-month follow-up interviews. For participants who were unable to return to the clinic, follow-up surveys were conducted via phone.

### Ethical approval

The study protocol was approved by the University of California San Diego’s Human Research Protections Program (Protocol 170084, February 7, 2017), the Population Council Institutional Review Board (Protocol 797, January 18, 2017) and the Kenyatta National Hospital-University of Nairobi Ethics and Research Committee (Protocol P945/12/2016, March 2, 2017). The protocol for this study has been published elsewhere.(17) All participants completed signed informed consent. For participants age 15 to 17 parental consent was waived as research was sought within the context of confidential reproductive health services in Kenya.

### Intervention

The ARCHES intervention was first developed and evaluated in the US.(18–20) Adaptation to the Kenyan context was conducted using qualitative interviews, cognitive interviewing, and iterative workshopping with local partners and providers.(13, 17) Details of the adaptation process are reported elsewhere.(17) Implementation of ARCHES includes training of providers who currently offer contraceptive counseling to include ARCHES elements in this form of care. Implementation is further facilitated via in-clinic mentoring and provision of feedback based on client exit surveys indicating changes in the quality of implementation over time(17).

ARCHES is designed to educate and support women’s agency to address IPV and RC to enable successful contraceptive use irrespective of family opposition, thereby reducing unintended pregnancy. Elements include provision of comprehensive contraceptive counseling and education, inclusive of education on RC and strategies to use contraceptive methods without partner knowledge universally regardless of RC disclosure. Following discussions aimed at destigmatizing RC and improving contraceptive self-efficacy, and building on trust established earlier in the visit, ARCHES providers screen all female patients with whom they can secure privacy for IPV and RC as a component of standard contraceptive counseling. Understanding that disclosure to providers is uncomfortable or frightening to some,(24–26) screening involves no pressure or coercion to disclose. Women who disclosure either form of abuse are supported with a rights-based response and further first-line response counseling. Women who disclose IPV specifically are offered the option to be connected by phone to a local gender-based violence support service at the conclusion of their family planning visit. No external referrals are offered for clients disclosing RC as the intervention content directly addresses this coercion, including support for implementing strategies to cope with this challenge to their reproductive autonomy (e.g. covert contraceptive use). At the completion of the ARCHES clinic visit, all clients are offered palm-sized booklets with information about IPV and RC, covert contraceptive methods, and contact information for local IPV support services. To minimize potential patient harm, ARCHES providers prioritize patient confidentiality, providing counseling only where privacy (visual and auditory) is ensured. In the absence of privacy, patients receive the local standard of care contraceptive counseling. Given development efforts and provider norms supporting male engagement in family planning in the Kenyan context (27, 28), the adaptation also included strategies on how to separate an accompanying male partner safely and, at the end of counseling, an offer for women to bring their male partner into the appointment for more information on contraceptive methods (but without pressure to do so).

### Measures

#### Primary outcomes

Primary distal outcomes of interest for this study are RC and physical and sexual IPV and measured at the individual participant level. All outcomes pertain to the individual participant level.

RC is as a binary measure, dichotomized using a ten-item scale adapted from a scale originally developed in the US(29) and refined by the study team using formative research and cognitive interviewing in Kenya on baseline and follow-up surveys. Six items were included on pregnancy coercion (“tried to force or pressure you to become pregnant”; “said he would leave you if you did not become pregnant”; “threatened to hurt or beat you if you tried to prevent or delay getting pregnant”; “told you he would have a baby with someone else if you didn’t get pregnant”; “said you could not use family planning because you do not have enough sons”; “insulted you, yelled at you, or made you feel badly for using or wanting to use family planning”), three on contraceptive sabotage (“intentionally broken a condom; poked holes in a condom or taken a condom off during sex to try to get you pregnant”; “stopped you from going to a clinic or community health event to obtain family planning”; “ever destroyed, hidden, or taken away your family planning method”), and two on abortion coercion (“pressured you to end a pregnancy when you did not want to end it”; “pressured you to keep a pregnancy that you wanted to end”). After each binary item a time question established the last time this action happened with the current partner. Those answering “within the past three-months” on any of the nine-scaled items were dichotomized yes for past three-month RC. A similar two-item screening measure was assessed for RC at exit asking “Have you ever felt pressured or forced by your current partner to become pregnant when you did not want to be?” and “Has your current partner ever made it difficult for you to get family planning or to use family planning (e.g. destroy, hide, or take away your contraception)?”. RC was measured at baseline, exit, three- and six-month follow-up. Baseline and exit measures were combined (if positive to RC at baseline or exit interview then positive for RC) due to differences in reporting of RC between timepoints and treatment groups and inability for RC exposure to change without leaving the clinic. Similarly, RC exposure at three- and six-month follow-up timepoints were combined (if positive for RC at three- or six-month follow-up then positive for RC in the past six-months) to increase power to detect effects for rare outcomes.

Physical IPV was assessed using adaptations of the Conflict Tactics Scale 2-Short form(30) dichotomized based on any yes response to six items (“ever pushed you, shaken you or thrown something at you”; “ever slapped you”, “ever twisted your arm or pulled your hair”, “ever hit you with his fist or something that could hurt you”, “ever kicked you, dragged you, or beaten you up”, “ever tried to choke or burn you”). Sexual IPV was assessed using one question adapted from the Sexual Experiences Short Form Survey(31) on baseline and follow-up surveys (“ever forced you to have sex or do something sexual with you when you didn’t want to”). These questions asked about lifetime experience. For those who responded yes to any of the physical or sexual IPV items, a follow-up question asked about the “last time that this happened to you.” Those answering “within the past three-months” on were dichotomized yes for IPV. Another screening question was included at exit for physical IPV, “Are you currently in a relationship with a partner who physically hurts you?” and sexual IPV, “Are you currently in a relationship with a partner who makes you have sex or do something sexual that makes you feel uncomfortable?” IPV was measured at baseline, exit, three- and six-month follow-up. Baseline and exit measures were combined as well as three- and six-month follow-up time points in the same manner and for the same reason as described for RC.

#### Secondary outcomes

Secondary distal outcomes of interest included leaving an abusive relationship, incident pregnancy and unintended pregnancy, measured at follow-ups only.

Leaving an abusive relationship was measured at three- and six-month follow-up using the question “In the last three months, have you left a relationship because it felt unhealthy, unsafe or abusive?”, adapted from similar measures in prior ARCHES trials.(18, 19)

Incident pregnancy was measured at three- and six-month follow-up by one question asking women how many times they had been pregnant in the prior three months, including miscarriages, abortions and stillbirths with those indicating one or more pregnancies dichotomized to experiencing pregnancy.

Unintended pregnancy in the past three months was assessed at three- and six-month follow-up using one question adapted from the National Survey of Family Growth(32) “At the time you became pregnant, did you want to become pregnant then, did you want to wait to become pregnant at a later date, or did you not want any more children?”. Unintended pregnancy was categorized as none in the past three months (not pregnant in past three months or wanted to become pregnant) vs. any in the past three months (became pregnant and wanted to become pregnant later or not at all). Like RC and IPV outcomes, leaving an abusive relationship, incident pregnancy and incident unintended pregnancy at three- and six-month follow-up timepoints were combined to increase power to detect effects for rare outcomes.

#### Covariates

Key patient-level covariates include age (continuous), marital status (married or cohabitating vs. unmarried), highest education level attended (primary or less vs. secondary vs. tertiary or higher), parity (nulliparous vs. uniparous vs. multiparous), recent employment (paid work in past 12 months vs. no paid work in past 12 months), past 30-day food insecurity (any family members went without heating the whole day because there was not enough food vs. no family members went hungry for a whole day), and current contraceptive use (current modern contraceptive use vs. no current modern contraceptive use). Covariates were selected based on a-priori hypotheses of being potential confounders and presence of differences between treatment groups at baseline.

### Sample size

Study sample size was determined by 80% power to detect an eight-percentage point reduction in intervention group reporting (equivalent to a 0.4 reduction in odds) of RC with an intra-class correlation of <0·1 based on prior ARCHES study effect sizes.^23^ Calculations assumed three intervention and three control clusters (six total), unequal cluster sizes ranging from approximately 50 to 250 clients with a mean cluster size of 100, and a coefficient of variation for cluster size of 0·5. Baseline prevalence of RC was estimated at 17·14 percent based on preliminary results from the pilot. Under these parameters, the required average cluster size was 68 participants, yielding a total analytic sample of 408 clients (204 per arm) retained at the final follow-up. Baseline RC prevalence was estimated at 17·14% from pilot data among 143 FHOK clients ages 15–49 years. Anticipating retention of 85% at three months and 75% at six months—based on prior ARCHES retention patterns—we calculated a minimum baseline sample of 544 participants.

### Analyses

Primary analyses comprised of a modified intent-to-treat (mITT) approach.(33) All patients who reported receiving any contraceptive counseling on their exit survey were included in the analytical sample and included in the treatment arm (intervention/control) assigned to the clinic they attended. mITT was employed over a standard ITT approach, which would have conducted analyses on all participants enrolled at baseline regardless of reporting receipt of contraceptive counseling at exit, as over 15% of patients in the control group who completed baseline received no contraceptive counseling at their appointment. This is despite those individuals indicating that they were interested in receiving contraceptive counseling at their appointment at the time of enrollment.

Secondary analyses employed an as-treated approach; from the mITT sample, patients from the intervention group were moved to the control group if they did not report receiving all of the four core ARCHES strategies as the intervention intended (i.e. screening for RC, IPV, information on methods that can be used discretely and referral information) at exit. To further clarify, participants from the intervention group reporting receipt of some or none of the four core intervention strategies were reclassified as control group participants and compared to those in the intervention group who received the full intervention.

Descriptive statistics were conducted to look at prevalence of all measures at baseline. Descriptive statistics and bivariate hypothesis tests, Pearson’s Chi-squared for binary and categorical covariates and Satterthwaite T-test for unequal variances for continuous covariates, also tested if those retained through six month follow-up were different than those who were loss-to-follow-up at both three and six-month follow-ups. The intervention and control groups were significantly different on multiple covariates assessed at baseline including age, marital status, education, parity, employment food insecurity and current contraceptive use.). To account for this, all covariates were used to create inverse probability of treatment weights (IPTW) to correct group imbalances.(34) IPTW balanced all covariates between mITT treatment and control groups so that there were no significant differences at alpha <·05 (Table 1). IPTW weights utilized ATT (average treatment effect on the treated) weighting, thus, control observations were weighted to balance with original treatment observations (unweighted).(35) ATT weighting was employed because the treatment group reported higher rates of RC and IPV at baseline (Table 1) suggesting greater vulnerability to violence than controls. The same strategy was also applied for the as-treated analyses, where IPTW similarly balanced treatment groups (Supplement 1).

**Table 1.**
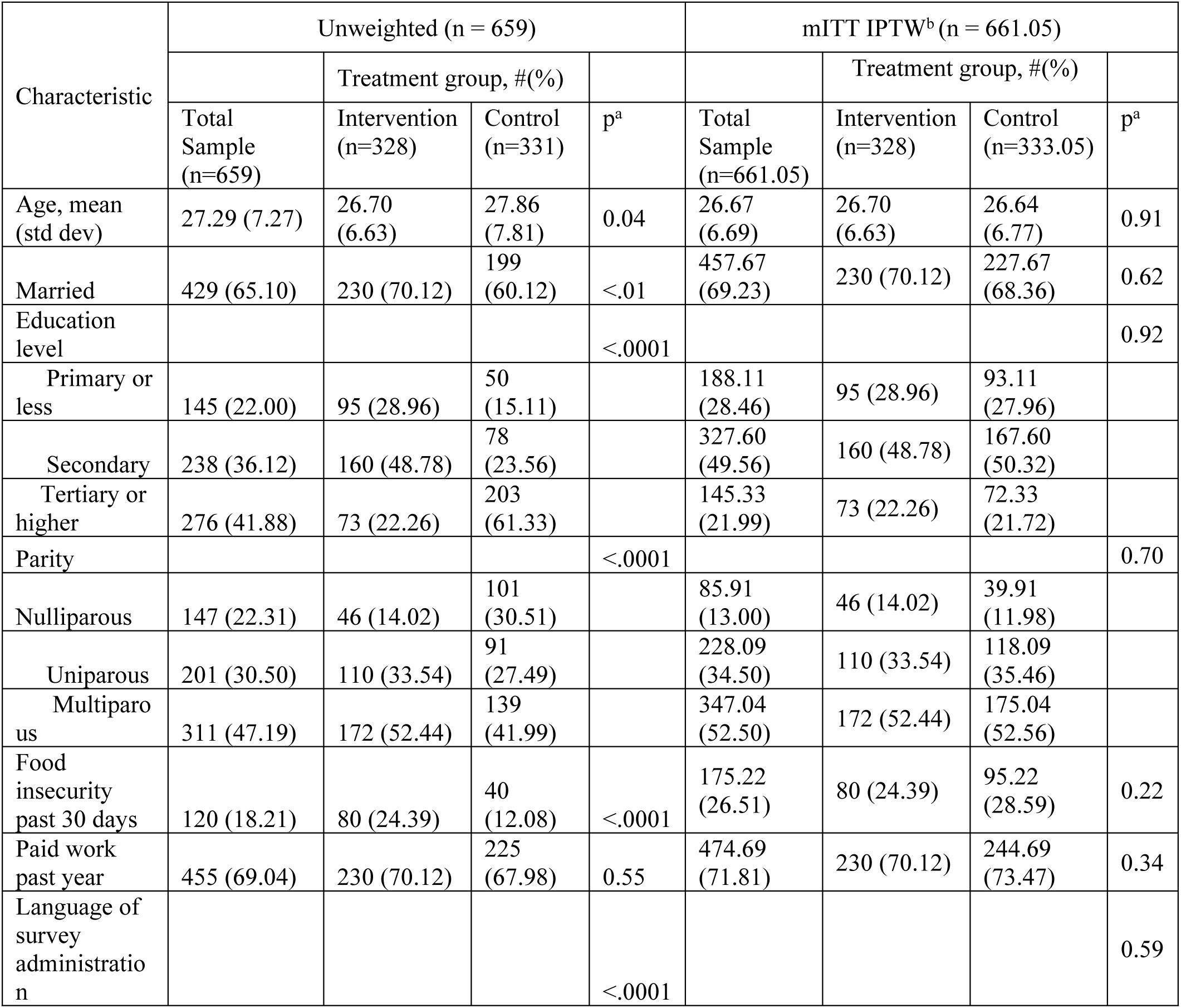
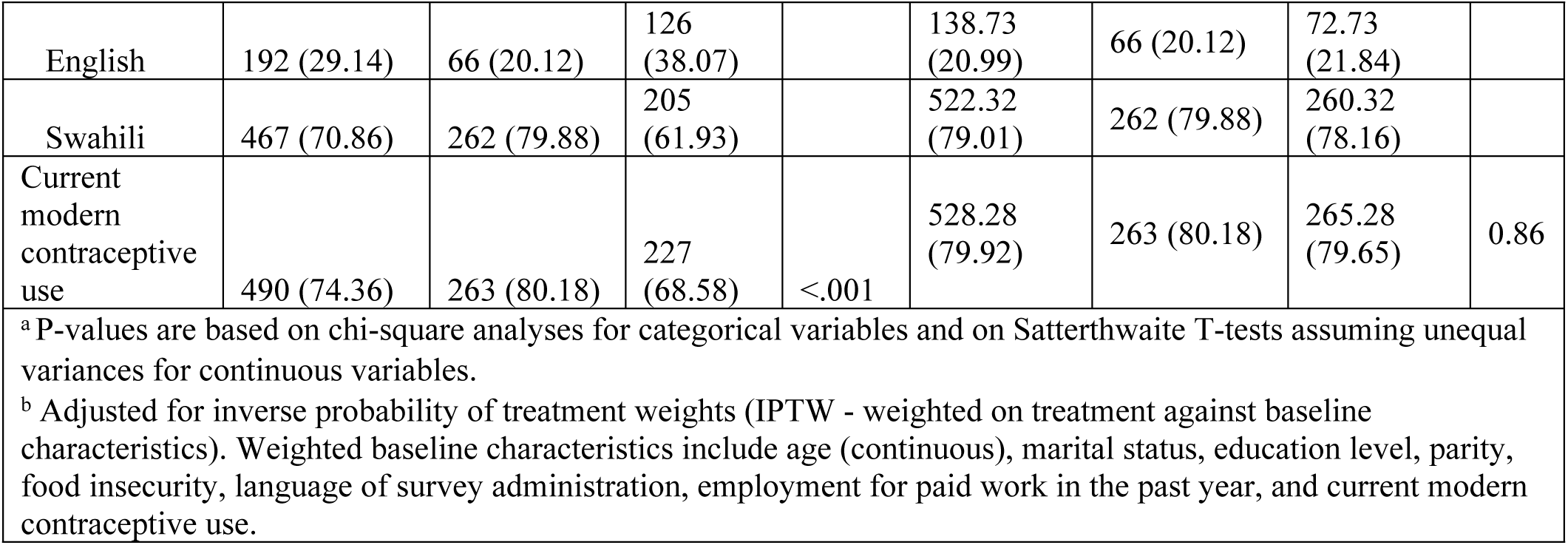
Female family planning client characteristics at baseline by ARCHES modified intent-to-treat (mITT) treatment groups, unweighted (unbalanced) and IPTW (balanced) (Nairobi, Kenya; 2019).

| Characteristic | Unweighted (n = 659) |  |  |  | mITT IPTW<sup>b</sup> (n = 661.05) |  |  |  |
| --- | --- | --- | --- | --- | --- | --- | --- | --- |
|  |  | Treatment group, #(% ) |  |  |  | Treatment group, #(% ) |  |  |
|  | Total Sample (n=659) | Intervention (n=328) | Control (n=331) | p<sup>a</sup> | Total Sample (n=661.05) | Intervention (n=328) | Control (n=333.05) | p<sup>a</sup> |
| Age, mean (std dev) | 27.29 (7.27) | 26.70 (6.63) | 27.86 (7.81) | 0.04 | 26.67 (6.69) | 26.70 (6.63) | 26.64 (6.77) | 0.91 |
| Married | 429 (65.10) | 230 (70.12) | 199 (60.12) | <.01 | 457.67 (69.23) | 230 (70.12) | 227.67 (68.36) | 0.62 |
| Education level |  |  |  | <.0001 |  |  |  | 0.92 |
| Primary or less | 145 (22.00) | 95 (28.96) | 50 (15.11) |  | 188.11 (28.46) | 95 (28.96) | 93.11 (27.96) |  |
| Secondary | 238 (36.12) | 160 (48.78) | 78 (23.56) |  | 327.60 (49.56) | 160 (48.78) | 167.60 (50.32) |  |
| Tertiary or higher | 276 (41.88) | 73 (22.26) | 203 (61.33) |  | 145.33 (21.99) | 73 (22.26) | 72.33 (21.72) |  |
| Parity |  |  |  | <.0001 |  |  |  | 0.70 |
| Nulliparous | 147 (22.31) | 46 (14.02) | 101 (30.51) |  | 85.91 (13.00) | 46 (14.02) | 39.91 (11.98) |  |
| Uniparous | 201 (30.50) | 110 (33.54) | 91 (27.49) |  | 228.09 (34.50) | 110 (33.54) | 118.09 (35.46) |  |
| Multiparous | 311 (47.19) | 172 (52.44) | 139 (41.99) |  | 347.04 (52.50) | 172 (52.44) | 175.04 (52.56) |  |
| Food insecurity past 30 days | 120 (18.21) | 80 (24.39) | 40 (12.08) | <.0001 | 175.22 (26.51) | 80 (24.39) | 95.22 (28.59) | 0.22 |
| Paid work past year | 455 (69.04) | 230 (70.12) | 225 (67.98) | 0.55 | 474.69 (71.81) | 230 (70.12) | 244.69 (73.47) | 0.34 |
| Language of survey administration |  |  |  | <.0001 |  |  |  | 0.59 |
| English | 192 (29.14) | 66 (20.12) | 126 (38.07) |  | 138.73 (20.99) | 66 (20.12) | 72.73 (21.84) |  |
| Swahili | 467 (70.86) | 262 (79.88) | 205 (61.93) |  | 522.32 (79.01) | 262 (79.88) | 260.32 (78.16) |  |
| Current modern contraceptive use | 490 (74.36) | 263 (80.18) | 227 (68.58) | <.001 | 528.28 (79.92) | 263 (80.18) | 265.28 (79.65) | 0.86 |
| <sup>a</sup> P-values are based on chi-square analyses for categorical variables and on Satterthwaite T-tests assuming unequal variances for continuous variables.<br><sup>b</sup> Adjusted for inverse probability of treatment weights (IPTW - weighted on treatment against baseline characteristics). Weighted baseline characteristics include age (continuous), marital status, education level, parity, food insecurity, language of survey administration, employment for paid work in the past year, and current modern contraceptive use. |  |  |  |  |  |  |  |  |

ARCHES’ effects on RC, sexual and physical IPV were measured using difference-in-differences (DiD) multivariable, mixed-effects logistic IPTW models. Fixed effects, for analyses assessing the change over time, included parameters for time, treatment and time by treatment. Time comprised of baseline (0) and combined three- and six-month follow-up (1). Random effects included parameters for within variance of clinic-clusters and repeated observations of individuals nested within clusters over time. The interaction term (time by treatment) beta was interpreted to estimate the average treatment effect (i.e., the relative change over time between treatment groups or the difference between slopes of treatment groups). From DiD models, we also compared treatment groups at each timepoint (baseline and follow-up) using estimated odds ratios (holding time point constant and comparing treatment groups from the time by treatment interaction term) to aid in interpretation. DiD was applied with IPTW given that treatment groups were highly unbalanced across primary outcomes and covariates at baseline, an approach which has proven effective in estimating unbiased treatment effects in other non-randomized intervention studies.(36, 37)

Differences between intervention and control groups at follow-up for secondary outcomes of leaving an abusive relationship, incident pregnancy, and unintended pregnancy were measured using mixed-effects single timepoint logistic regression models. The fixed effect included treatment and random effect accounted for within variance of clinic-clusters. The effect of treatment among the treated was interpreted from the exponentiated treatment parameter (i.e. odds ratio). DiD estimators could not be fit for secondary outcomes as they were not assessed at baseline.

We conducted a post-hoc exploratory analysis to assess whether the intervention’s effect on unintended pregnancy may be partially explained by contraceptive uptake at exit, a significant proximal outcome (23). We re-estimated the primary models including contraceptive uptake (yes/no) at follow-up as an additional covariate to evaluate any attenuation of the intervention effect. This approach provides descriptive insight but does not meet assumptions required for causal mediation and therefore should not be interpreted causally.

All analyses were conducted using SAS v. 9·4 (SAS Institute Inc, Cary, NC).

### Reflexivity and positionality statement

The lead author is a white, US-based male senior professor student with over three decades of experience in sexual and reproductive health and rights and gender-based violence prevention intervention evaluations. The authorship team includes both male and female individuals at various career stages ranging from graduate trainees to mid-career professionals and senior researchers. The team consists of US-based, UK-based, and Kenyan nationals. We acknowledge the influence of our positionalities and engaged per protocol reporting and analysis and in reflexive discussions to interpret quantitative results to mitigate reporting biases.

## Results

### Sample description

Three clinics were allocated to intervention and control, each. 3,678 clients were assessed for eligibility (1315 intervention; 2363 control) and 2,945 were excluded (972 intervention; 1973 control), mostly due to ineligibility. A total of 29 clients declined to participate. In total, 659 women and girls ages 15-49 were enrolled in ARCHES and completed the exit survey (n=328 intervention, n=331 control) between July 2018 and December 2018 (Figure 1). Originally, 733 clients were enrolled in baseline, but 74 of these (15 intervention; 59 control) were excluded as they reported not receiving any FP services or counseling. The final sample resulted in 659 clients mITT (mITT: 328 intervention, 331 control. Within the treatment group, 77% of participants reporting receiving the core ARCHES strategies at exit and were allocated to the intervention group for as-treated analyses (as-treated: 254 intervention, 405 control, Supplement 1). From the eligible sample, six-month retention was 87% in the intervention arm, and 80% in the control arm. There were no significant differences between those retained and those loss-to-follow-up at both three and six-month follow-up on demographic variables or rates of RC, IPV, or SV at baseline (Supplement 2). The trial commenced when the sample size was reached and all enrolled participants had been contacted to complete baseline.

**Figure 1.**
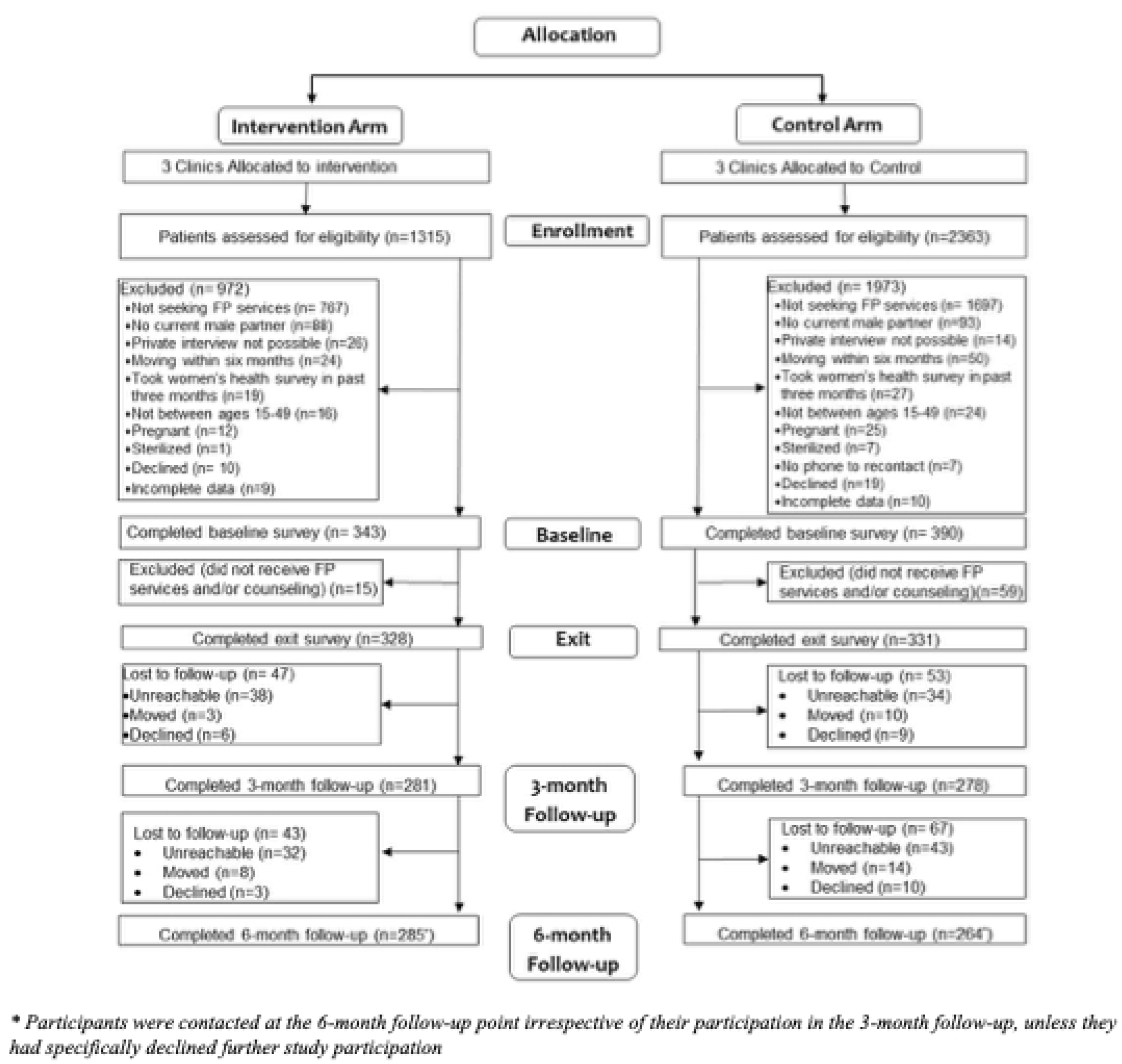
ARCHES cluster-controlled trial consort flow chart (Nairobi, Kenya; 2019).

Participants were an average of 27 years old, and were majority married (65%), current contraceptive users (90%) and food secure (82%) (Table 1). More than one in three participants had secondary level education (36%), and more than one in five had achieved tertiary or higher education (22%). Nearly half of participants had two or more children (47%), and 22% had not yet given birth. Women enrolled in the ARCHES intervention arm in the mITT analysis were younger, less educated, had more children, had higher food insecurity, and were more likely to be using contraceptives and be married than women enrolled in the control arm. Somewhat similarly, in the as-treated analysis, women receiving the full intervention were younger, less educated, had more children, had lower food insecurity, and were more likely to be using contraceptives and be married than women in the control arm (Supplement 1). Differences in sociodemographics were neutralized by the IPTW procedure for both mITT and as-treated data sets, which generated balanced datasets across intervention and control arms (Table 1, Supplement 1).

### ARCHES Effects on Primary Distal Outcomes

At baseline, levels of RC were higher among the ARCHES arm than the control arm (26% vs. 18%, Table 2, Figure 2; AOR 2·00, 95% CI 1·25, 3·22, p=0.004, Supplement 3). This difference increased at follow-up, where RC among ARCHES participants was 18% vs. 6% in the control group (OR 5·96, 95% CI 4·43, 8·02, p<0·001, Supplement 3, Figure 2). Both treatment groups decreased rates of RC between baseline and follow-up based on the mITT approach (control 12%, p<·001; intervention 8%, p=0·022, Table 2). In the as-treated approach, the reduction in RC among ARCHES participants was not as substantial. Both mITT and as-treated DiD models indicated that the control group experienced larger reductions than the intervention group, yielding effect estimates that appeared as relative increases in RC among those receiving the ARCHES intervention (mITT time x treatment beta = 1·09, 95% CI 0·81, 1·37, p<·001, clinic-level ICC = 0·009; as-treated time x treatment beta = 1·08, 95% CI 0·87, 1·30, p<·001, clinic-level ICC = 0·069, Table 3).

**Table 2.** IPTW descriptive outcomes at each time point within treatment groups (Nairobi, Kenya; 2019).

|  | Control (weighted) |  |  | ARCHES Intervention |  |  |
| --- | --- | --- | --- | --- | --- | --- |
|  | Baseline | 6-month follow-up |  | Baseline | 6-month follow-up |  |
|  | n (%) | n (%) | p<sup>b</sup> | n (%) | n (%) | p<sup>b</sup> |
| ***mITT approach*** | n=333.05 |  |  | n=327 |  |  |
| Reproductive coercion (past 3 mo) | 59.41 (17.91) | 16.51 (5.56) | <.0001 | 84 (25.69) | 53 (18.03) | 0.02 |
| Physical IPV (past 3 mo) | 40.59 (12.28) | 46.97 (15.80) | 0.20 | 76 (23.31) | 41 (13.90) | <.01 |
| Sexual IPV (past 3 mo) | 33.60 (10.09) | 12.52 (4.21) | <.01 | 59 (17.99) | 46 (15.59) | 0.43 |
| Leaving an abusive relationship | - | 51.75 (17.41) | - | - | 61 (20.68) | - |
| Incident pregnancy | - | 25.36 (8.54) | - | - | 11 (3.78) | - |
| Incident unintended pregnancy | - | 10.47 (3.52) | - | - | 5 (1.72) | - |
| **As-treated approach** | n=256.66 |  |  | n=254 |  |  |
| Reproductive coercion (past 3 mo) | 58.82 (23.00) | 17.74 (8.05) | <.0001 | 57 (22.53) | 43 (18.61) | 0.29 |
| Physical IPV (past 3 mo) | 41.91 (16.43) | 32.95 (14.94) | 0.66 | 61 (24.21) | 34 (14.66) | <.01 |
| Sexual IPV (past 3 mo) | 35.60 (13.88) | 21.59 (9.79) | 0.17 | 45 (17.72) | 31 (13.36) | 0.19 |
| Leaving an abusive relationship | - | 33.40 (15.15) | - | - | 53 (22.84) | - |
| Incident pregnancy | - | 18.64 (8.50) | - | - | 6 (2.62) | - |
| Incident unintended pregnancy | - | 7.03 (3.20) | - | - | 3 (1.32) | - |
| <sup>b</sup> P-values are based on chi-square analyses. |  |  |  |  |  |  |

**Figure 2.**
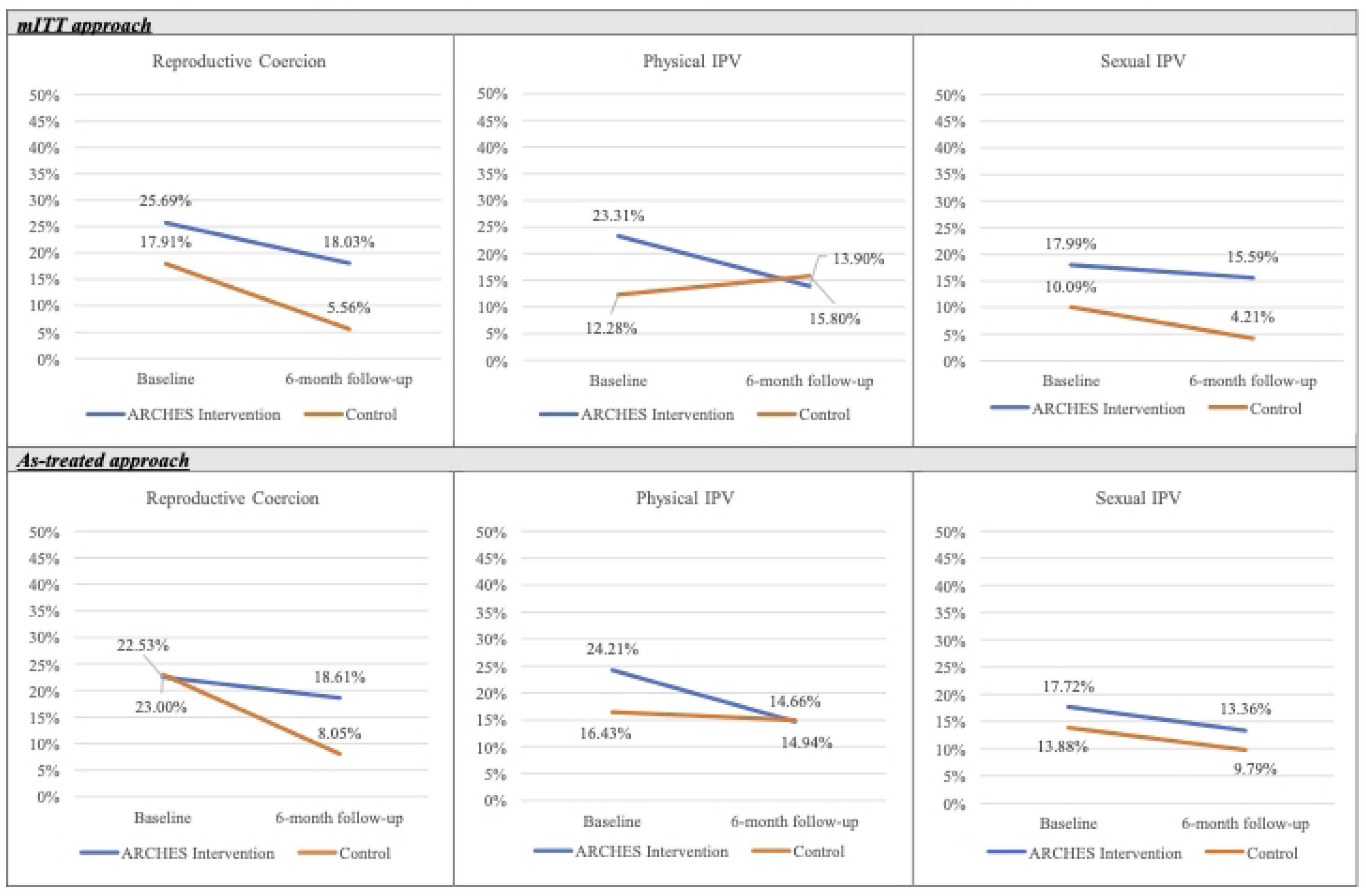
IPTW descriptive outcomes [proportions (binary) and means (continuous)] graphed over time within treatment groups (Nairobi, Kenya; 2019)

**Table 3.** Difference-in-differences average treatment effect among the treated (time x treatment) comparing treatment groups (ref = control) from multi-variable mixed-effect logistic regression (Nairobi, Kenya; 2019).

| Outcome | mITT IPTW time x treatment beta (95% confidence interval)<sup>b</sup> | p-value | As-treated IPTW time x treatment beta (95% confidence interval)<sup>b</sup> | p-value |
| --- | --- | --- | --- | --- |
| Reproductive coercion | 1.09 (0.81, 1.37) | <.0001 | 1.08 (0.87, 1.30) | <.0001 |
| Physical intimate partner violence | -0.98 (-1.58, -0.39) | <0.01 | -0.56 (-1.19, 0.07) | 0.08 |
| Sexual intimate partner violence | 1.16 (0.47, 1.85) | <0.01 | 0.06 (-0.74, 0.85) | 0.89 |
| <sup>a</sup> Adjusted for inverse probability of treatment weights (weighted on treatment against baseline characteristics), fixed effects (time, treatment, time x treatment) and random effects (cluster and individual nested with cluster). Coefficient shown is the exponentiated time x treatment interaction term. |  |  |  |  |

Women in the ARCHES intervention group reported higher levels of pIPV at baseline as compared to controls (23% vs. 12%, Figure 2; AOR 2·98, 95% CI 1·62, 5·46, p<·001, Supplement 3) but by follow-up this difference was smaller (14% vs. 16%, Figure 2; AOR 1·11, 95% CI 0·42, 2·95, p=0·827, Supplement 2). Within group mITT and as-treated analyses show significant decreases in physical IPV over time for the ARCHES intervention group, while control participants saw a slight increase in increase in reports of physical IPV (Table 2). DiD models found a greater relative reduction in physical IPV based on receiving the ARCHES intervention via the mITT approach (time x treatment beta = -0·98, 95% CI -1·58, -0·39, p=0·001, clinic-level ICC = 0·044) and a reduction via the as-treated approach (time x treatment beta = -0·56, 95% CI -1·19, 0·07, p=0·083, clinic-level ICC = 0·050, Table 3) as compared to controls.

Similarly, women attending ARCHES clinics reported higher levels of sexual IPV compared to women attending control clinics at baseline (18% vs. 10%, Figure 2; AOR 2·66, 95% CI 1·69, 4·18, p<·001, Supplement 3). While sexual IPV reports decreased at follow-up in both groups (16% intervention vs. 4% control, Figure 2) this decrease was only significant among the control group for the mITT approach (Table 2). DiD models indicated that the control group experienced a larger reduction than the intervention group, yielding an mITT effect estimate that appeared as a relative increase in sexual IPV among those receiving the ARCHES intervention (time x treatment beta = 1·16, 95% CI 0·47, 1·85, p<·001, clinic-level ICC = 0·002). Results differed for the as-treated approach, which over time showed a parallel time trend between treatment groups (Figure 2, time x treatment beta 0·06, 95% CI -0·74, 0·85, p=0·889, clinic-level ICC = 0·077).

### ARCHES Effects on Secondary Distal Outcomes

Although a larger percentage of women in the ARCHES group reported leaving an abusive relationship at follow-up than those in the control group (23% vs. 15%, Table 2), this difference was only significant in as-treated adjusted models (mITT AOR 1·40, 95% CI 0·58, 3·42, p=0·457, clinic-level ICC =0·103 ; as-treated AOR 2·14, 95% CI 1·14, 4·04, p=0·018, clinic-level ICC = 0·062, Table 4).

**Table 4.** Average intervention effect at follow-up comparing treatment groups (ref = control) from multi-variable mixed-effect logistic regression (Nairobi, Kenya; 2019).

| Outcome | mITT IPTW<br>odds ratio<br>(95% confidence<br>interval)<sup>a</sup> | p-value | As-treated IPTW<br>odds ratio<br>(95% confidence<br>interval)<sup>b</sup> | p-value |
| --- | --- | --- | --- | --- |
| Leaving an abusive<br>relationship | 1.40 (0.58, 3.42) | 0.46 | 2.14 (1.14, 4.04) | 0.02 |
| Incident pregnancy | 0.42 (0.27, 0.67) | <.001 | 0.29 (0.08, 1.02) | 0.05 |
| Unintended<br>pregnancy | 0.48 (0.16, 1.45) | 0.19 | 0.39 (0.09, 1.63) | 0.19 |
| <sup>a</sup> Adjusted for a fixed effect (treatment) and a random effect (cluster) only. Coefficient shown is for ARCHES participation (reference: control group).<br><sup>b</sup> Adjusted for inverse probability of treatment weights (weighted on treatment against baseline characteristics), a fixed effect (treatment) and a random effect (cluster). Coefficient shown is for ARCHES participation (reference: control group) |  |  |  |  |

The ARCHES intervention group was significantly less likely to report incident pregnancy during the follow-up period as compared to controls in both mITT and as-treated models (mITT 4% vs. 9%, AOR 0·42, 95% CI 0·27, 0·67, p<·001, clinic-level ICC = 0; as-treated 3% vs 9%, AOR 0·29, 95% CI 0·08, 1·02, p=·054, clinic-level ICC = 0, Table 4). When we accounted for contraceptive uptake in the models, we found that controlling for this proximal outcome did slightly attenuate the effect, shifting the mITT model to an AOR of 0·50, 95% CI 0·31, 0·83 and the as-treated model to an AOR 0·32, 95% CI 0·10, 1·04 (Supplement 4). Similarly, fewer women reported an unintended pregnancy during the follow-up period among the ARCHES intervention group at follow-up but this was not significant in either model (mITT 2% vs. 4%, AOR 0·48, 95% CI 0.16, 1.45, p=0.193, clinic-level ICC =0·056; as-treated 1% vs 3%, AOR 0·39, 95% CI 0.09, 1.63, p=0.195, clinic-level ICC = 0·026, Table 4).

## Discussion

The current study sought to assess the effects of the ARCHES intervention on the primary outcomes of RC and physical and sexual IPV and on the secondary outcomes of leaving an abusive relationship and incident and unintended pregnancy among women seeking contraceptive counseling in six community-based clinics in Nairobi, Kenya via a cluster-controlled trial. Results were mixed. As compared to the reduction in RC observed among controls, women receiving ARCHES reported a relative increase in exposure to this behavior. Receiving ARCHES vs. standard-of-care services was associated with reduced odds of physical IPV at follow-up. Similar to the finding for RC, reductions in sexual IPV related to receipt of ARCHES were smaller than those reported by control participants, demonstrating a relative increase in this form of abuse; this negative effect was not observed in as-treated analyses. Among this sample of women seeking contraceptive services, ARCHES increased the odds of leaving an abusive partner and reduced the odds of becoming pregnant in the subsequent six months. Although fewer unintended pregnancies were reported by those receiving ARCHES as compared to those receiving the standard-of-care, this difference was not statistically significant.

Although significant reductions in recent RC were seen within both intervention and control groups over time, the magnitude of the reduction reported at 6-months follow-up by participants receiving standard contraceptive counseling was greater than that reported by ARCHES participants via both the mITT and as-treated approaches. It is difficult to explain why controls reported such a precipitous drop in RC after receiving standard care. One possible explanation for this result relates to the content and mechanisms of the ARCHES intervention. It is possible that the observed effects of decreased incident pregnancy and/or increased covert contraceptive use may mediate the effect of ARCHES exposure and RC. For example, as RC is largely motivated in this context to promote pregnancy(13), the finding supporting the success of ARCHES in reducing the odds of pregnancy may mean that women remain vulnerable to RC. Similarly, covert contraceptive use may increase risk for RC by delaying pregnancy and/or through partner discovery of covert use. It is critical that future longitudinal research clarify whether and how resisting RC by such means as covert contraceptive use and pregnancy prevention affects future risk for coercion and other forms of violence. However, so as not to reduce women’s reproductive autonomy, findings of such research should not be used to narrow choices for contraceptive use presented to women (i.e., not supporting the option of covert use) but rather to inform women of potential risks associated with their choices. Additionally, learnings from this trial may support that, in this context, the intervention may not reduce RC behaviors enacted by male partners as we previously expected. Although a possible route for intervention-related direct effects on male partner RC perpetration was offered as requested in formative research (educational contraceptive counseling for male partners (13)), only 22 women (10 intervention, 12 control) chose to exercise this option (unpublished data) and only one of these women reported RC at baseline or follow-up. In contrast to this finding, a recent cluster RCT (cRCT) of ARCHES in Nigeria among a mixed sample of women seeking contraceptive care and antenatal care (ANC) found that ARCHES reduced the odds of RC at follow-up(22), while the recent cRCT of the intervention among women seeking abortion care in Bangladesh found no effect on RC(21). Whether and how ARCHES affects subsequent experiences of RC and how this differs across patient populations are critical topics for future research.

Significant reductions in physical IPV were seen among ARCHES participants (relative reduction of 40%) with no changes over time among controls via both mITT and as-treated approaches, yielding an intervention effect of reduced odds of such violence across models (although the effect in as-treated models was somewhat attenuated, with p=·083 for this model). The finding that receipt of ARCHES reduces women’s risk of physical IPV is also consistent with the content and mechanisms of this intervention. Specifically, ARCHES is designed to increase awareness of IPV and local services available to women experiencing IPV based on both the content of the counseling received and the palm-sized booklet provided that includes such information, and these expected effects were observed (18, 19, 23). In contrast with the mixed findings for RC across other ARCHES studies, both the Bangladesh and Nigeria trials found similar reductions in IPV(21, 22). Although neither of these studies segregated physical from sexual IPV, this consistent positive effect on partner violence across diverse geographic and care contexts indicates that the ARCHES model may be an important tool to address IPV within clinical care.

In a similar finding to that for RC, reductions in sexual IPV reported among ARCHES and control participants differed greatly for mITT (relative reduction of 13% vs. 58%) and as-treated (relative reduction of 25% vs. 29%) approaches, leading to a significant negative intervention effect in the mITT model and a null intervention effect in the as-treated model. As explained above, longitudinal studies further specifying mechanisms of ARCHES strategies including potential mediating relationships of incident pregnancy and covert contraceptive use on abuse outcomes are required to clarify these findings. An important takeaway from these findings is that the mechanisms and motivations behind physical IPV compared to sexual IPV and RC are likely different and future research should investigate motivations, mechanisms and effects on these forms of violence separately as targeted strategies may be required to successfully address and prevent different forms of abuse. Also as described above, neither previously published trial specified sexual IPV in their analyses, making comparisons difficult across trials. Given the current mixed findings for physical and sexual IPV, future ARCHES studies should include specification of differing forms of partner violence as well as mechanisms that may lead to reduction in some forms and not others.

Women receiving the ARCHES model of care were more likely than controls to report leaving an abusive relationship across the six-month follow-up period in as-treated models; this difference did not reach significance in ITT models of intervention effects. The same mechanisms described in the discussion of physical IPV are likely responsible for this finding, with qualitative interviews collected after the intervention trial among those reporting IPV at baseline and receiving ARCHES indicating that women used the palm-sized booklet to demonstrate to their partner that there were people who could help her and places that she could go if his abuse continued (forthcoming). In the Bangladesh trial, ARCHES was not found to impact the odds of this outcome(21); leaving an abusive relationship was not assessed in the Nigeria trial (22). One plausible explanation offered by the authors was the relatively high cost to women, the majority of whom were married, of leaving a male partner in the Bangladesh context (21).

As discussed above, ARCHES was found to reduce the risk of incident pregnancy at six months among this sample of Kenyan women seeking contraceptive care, and a descriptively lower number of women reported incident unintended pregnancy among ARCHES recipients vs. those receiving standard contraceptive services (p>.05). These results indicate that the ARCHES intervention may be a promising model to support women in their attempts to prevent pregnancy via clinic-based contraceptive counseling, though larger studies powered to detect this effect on unintended pregnancy are required to confirm this finding. Post-hoc analyses support the model’s theory of change that counseling supporting reproductive autonomy increased contraceptive uptake at exit (23), which partially contributed to reductions in incident pregnancy at follow-up, though increased contraceptive use at exit did not fully account for the effect on incident pregnancy. Other mechanisms of reductions in incident pregnancy, such as increased contraceptive control or agency, should be explored in future studies. The recent ARCHES trial in Bangladesh also found a reduction in incident pregnancy relative to controls (21). This outcome was not assessed in the Nigeria trial based on inclusion women receiving ANC (22). As both of these trials included participants either seeking to prevent or terminate a pregnancy, the consistent finding of reduced incident pregnancy indicates the likely utility of the ARCHES approach for supporting women’s reproductive agency.

### Limitations

In this study, despite an extensive adaptation process to modify the intervention to improve fidelity in a low resource context, high-quality implementation was a challenge, with almost one-in-four clients receiving care at an ARCHES clinic did not receive the intervention as intended. Qualitative research from this study highlights that while those received ARCHES had overwhelming positive reactions, some providers, particularly male providers, may have been hesitant or resistant to some aspects of ARCHES including counseling women regarding discreet use of contraceptive methods (i.e., without knowledge of their male partner) and supporting them to independently decide if they involved their partner or not in decisions related to contraceptive use, likely impacting implementation fidelity. Time constraints likely also impacted fidelity, with providers facing high patient volumes making the decision to skip all or certain aspects of ARCHES to allow them to meet the demand for care. Barriers to IPV referral and low client acceptability of IPV referral may also have increased providers hesitancy to screen for IPV in some situations (38). In both the current and Nigeria ARCHES trials(38, 39), providers and clients reported that integration of RC and IPV elements into counseling was largely feasible and acceptable, but that it often required several weeks or months for these practices to become routine. It may be that the current trial in Nairobi provided insufficient time to reach high-quality implementation, biasing results towards the null. Future ARCHES studies should ensure that sufficient time and tracking of implementation are provided to reach faithful implementation prior to evaluation assessments, inclusive of identifying and addressing barriers to fidelity. Another important limitation potentially affecting the results of the current study is the lack of equivalence at baseline across intervention groups, specifically that women attending intervention clinics reported higher rates RC and physical and sexual IPV. These differences persisted even after weighting to reach equivalence regarding demographics via IPTW. This is a likely indicator of residual confounding, with women attending intervention clinics appearing to live in contexts within which these forms of violence are more likely. For example, exposure to ARCHES may affect women living in contexts with a high acceptability of violence differently than those living in context where such violence is less accepted. The nature of any such distortion in effects based on residual confounding is not possible to assess based on the available data, i.e., this lack of equivalence may have biased results towards or away from the null, or not at all, and the current results do not indicate a clear pattern in this regard. Reasons for this lack of equivalence likely include the small number of clinics eligible for inclusion (six) and the high variability in the contexts in which these clinics were situated (e.g., slum vs. suburb vs. upscale urban neighborhood). Although administrative records were utilized to balance groups across patient volume and demographics, these records were likely inaccurate. Another factor that may have affected the current results is the heightened awareness of RC and IPV brought about via the intervention(23), as this may have led to differential reporting at follow-up beyond any intervention-related changes in actual violence experiences. Relatedly, all data were self-report, and therefore subject to recall and social desirability biases. Another possible limitation was some of our outcomes of interest, particularly past 3-month sexual IPV and unintended pregnancy, were quite rare and necessitated the combining of 3- and 6-month follow up windows. Finally, the current findings cannot be generalized beyond the current population of women seeking contraceptive care outside of this small number of NGO-run facilities in Nairobi, Kenya.

## Conclusions

Despite these limitations, this controlled trial of an intervention to jointly address RC and reduce IPV in an LMIC context provides preliminary support for the potential effectiveness of such an approach to improving women’s safety and reproductive agency in this setting in that receipt of the ARCHES intervention resulted in reductions of physical IPV and incident pregnancy. Further research is required to understand negative results related to reproductive coercion and sexual IPV that may stem from women enacting strategies supported via ARCHES to prevent pregnancy (e.g., covert contraceptive use).

## Declarations

### Ethics approval and consent to participate

The study protocol was approved by the University of California San Diego’s Human Research Protections Program (Protocol 170084), the Population Council Institutional Review Board (Protocol 797) and the Kenyatta National Hospital-University of Nairobi Ethics and Research Committee (Protocol P945/12/2016). All participants in this study completed signed informed consent. The protocol for this study has been published elsewhere.(17)

### Consent for publication

All participants consented to deidentified data being published in aggregate form as part of signed informed consent. No other individual data is reported in this manuscript.

### Availability of data and materials

The datasets generated and/or analyzed during the current study are available in the Dryad repository, https://doi.org/10.6076/D1Q30G

### Author contributions

JGS, JU, SB, conceived of the research plan with assistance from CU, WL, and SW. Data collection was facilitated by WL and CU with remote coordination assistance from JGS and JU. The analytical approach was developed by JGS and JU with input from SB, EP, LM and NJ. Analyses were conducted by JU. All authors contributed to interpretation of the results. LM, JGS, and JU led drafting of the manuscript with input from all other authors.

## Acknowledgments

We would like to acknowledge the Emilie Schwarz for her assistance in reviewing the document according to the CONSORT extension guidelines for cluster randomized trials.

## Supporting Information

**S1 Table. Female family planning client characteristics at baseline by ARCHES as-treated treatment groups, unweighted (unbalanced) and IPTW (balanced) (Nairobi, Kenya; 2019).**

**S2 Table. Differences in outcomes between treatment groups at each time point from multi-variable mixed-effect logistic regression (Nairobi, Kenya; 2019).**

## References

1. Starrs AM, Ezeh AC, Barker G, Basu A, Bertrand JT, Blum R, et al. Accelerate progress-sexual and reproductive health and rights for all: report of the Guttmacher-Lancet Commission. Lancet. 2018;391(10140):2642–92.

2. Coll CVN, Ewerling F, García-Moreno C, Hellwig F, Barros AJD. Intimate partner violence in 46 low-income and middle-income countries: an appraisal of the most vulnerable groups of women using national health surveys. BMJ Global Health. 2020;5(1):e002208.

3. Cripe SM, Sanchez SE, Perales MT, Lam N, Garcia P, Williams MA. Association of intimate partner physical and sexual violence with unintended pregnancy among pregnant women in Peru. Int J Gynaecol Obstet. 2008;100(2):104–8.

4. Dasgupta A, Saggurti N, Ghule M, Reed E, Donta B, Battala M, et al. Associations between intimate partner violence and married women’s condom and other contraceptive use in rural India. Sex Health. 2018;15(5):381–8.

5. Maxwell L, Devries K, Zionts D, Alhusen JL, Campbell J. Estimating the effect of intimate partner violence on women’s use of contraception: a systematic review and meta-analysis. PLoS One. 2015;10(2):e0118234.

6. Maxwell L, Nandi A, Benedetti A, Devries K, Wagman J, García-Moreno C. Intimate partner violence and pregnancy spacing: results from a meta-analysis of individual participant time-to-event data from 29 low-and-middle-income countries. BMJ global health. 2018;3(1):e000304-e.

7. McDougal L, Silverman JG, Singh A, Raj A. Exploring the relationship between spousal violence during pregnancy and subsequent postpartum spacing contraception among first-time mothers in India. EClinicalMedicine. 2020;23:100414.

8. Silverman JG, Gupta J, Decker MR, Kapur N, Raj A. Intimate partner violence and unwanted pregnancy, miscarriage, induced abortion, and stillbirth among a national sample of Bangladeshi women. Bjog. 2007;114(10):1246–52.

9. WHO, on behalf of the United Nations Inter-Agency Working Group on Violence Against Women Estimation and Data (VAW-IAWGED). Violence Against Women Prevalence Estimates, 2018. Geneva: World Health Organization; 2021. Contract No.: 978 92 4 002225 6.

10. Silverman JG, Raj A. Intimate partner violence and reproductive coercion: global barriers to women’s reproductive control. PLoS Med. 2014;11(9):e1001723.

11. Wood SN, Dozier JL, Karp C, Desta S, Decker MR, Shiferaw S, et al. Pregnancy coercion, correlates, and associated modern contraceptive use within a nationally representative sample of Ethiopian women. Sex Reprod Health Matters. 2022;30(1):2139891.

12. Wood SN, Kennedy SR, Akumu I, Tallam C, Asira B, Zimmerman LA, et al. Understanding the Link Between Reproductive Coercion and Covert Use of Contraception as a Safety Strategy for Women Experiencing Violence in Nairobi’s Urban Informal Settlements. Violence Against Women. 2022:10778012221108422.

13. Boyce SC, Uysal J, DeLong SM, Carter N, Undie CC, Liambila W, et al. Women’s and girls’ experiences of reproductive coercion and opportunities for intervention in family planning clinics in Nairobi, Kenya: a qualitative study. Reprod Health. 2020;17(1):96.

14. Grace KT, Anderson JC. Reproductive Coercion: A Systematic Review. Trauma Violence Abuse. 2018;19(4):371–90.

15. Mankelkl G, Kassaw AB, Kinfe B. Factors associated with modern contraceptive utilization among reproductive age women in Kenya; evidenced by the 2022 Kenyan demographic and health survey. Contracept Reprod Med. 2024;9(1):10.

16. World Health Organization. Responding to intimate partner violence and sexual violence against women: WHO clinical and policy guidelines. Geneva, Switzerland: WHO; 2013. Contract No.: 978 92 4 154859 5.

17. Uysal J, Carter N, Johns N, Boyce S, Liambila W, Undie CC, et al. Protocol for a matched-pair cluster control trial of ARCHES (Addressing Reproductive Coercion in Health Settings) among women and girls seeking contraceptive services from community-based clinics in Nairobi, Kenya. Reprod Health. 2020;17(1):77.

18. Miller E, Decker MR, McCauley HL, Tancredi DJ, Levenson RR, Waldman J, et al. A family planning clinic partner violence intervention to reduce risk associated with reproductive coercion. Contraception. 2011;83(3):274–80.

19. Miller E, Tancredi DJ, Decker MR, McCauley HL, Jones KA, Anderson H, et al. A family planning clinic-based intervention to address reproductive coercion: a cluster randomized controlled trial. Contraception. 2016;94(1):58–67.

20. Tancredi DJ, Silverman JG, Decker MR, McCauley HL, Anderson HA, Jones KA, et al. Cluster randomized controlled trial protocol: addressing reproductive coercion in health settings (ARCHES). BMC Womens Health. 2015;15:57.

21. Pearson E, Paul D, Menzel J, Shakhider MAH, Konika RA, Uysal J, et al. Effectiveness of the Addressing Reproductive Coercion in Health Settings (ARCHES) intervention among abortion clients in Bangladesh: a cluster-randomized controlled trial. EClinicalMedicine. 2024;73:102699.

22. Betron M, Bryce, E., Obafemi, S., Yusuf, S., Abdullahi, H., Ifemenam, E., Dikeocha, N., Maiakwai, A., Kabir, B., Oduenyi, C. Effectiveness of a clinic-based counselling intervention on risk of experiencing intimate partner violence and reproductive coercion: a matched-pair cluster-controlled trial in Ebonyi and Sokoto, Nigeria. BMJ Global Health. 2025;10.

23. Uysal J, Boyce SC, Undie CC, Liambila W, Wendoh S, Pearson E, et al. Effects of a clinic-based reproductive empowerment intervention on proximal outcomes of contraceptive use, self-efficacy, attitudes, and awareness and use of survivor services: a cluster-controlled trial in Nairobi, Kenya. Sex Reprod Health Matters. 2023;31(1):2227371.

24. Katiti V, Sigalla GN, Rogathi J, Manongi R, Mushi D. Factors influencing disclosure among women experiencing intimate partner violence during pregnancy in Moshi Municipality, Tanzania. BMC Public Health. 2016;15:715.

25. Boudreau CL, Kress H, Rochat RW, Yount KM. Correlates of disclosure of sexual violence among Kenyan youth. Child abuse & neglect. 2018;79:164–72.

26. Shah PS, Shah J. Maternal exposure to domestic violence and pregnancy and birth outcomes: a systematic review and meta-analyses. Journal of women’s health. 2010;19(11):2017–31.

27. FP2030. FP2030: Kenya 2024 [Available from: https://www.fp2030.org/kenya/.

28. Kenya Ministry of Health. Family Planning and Reproductive Health: Where are the Men? ; 2015.

29. McCauley HL, Silverman JG, Jones KA, Tancredi DJ, Decker MR, McCormick MC, et al. Psychometric properties and refinement of the Reproductive Coercion Scale. Contraception. 2017;95(3):292–8.

30. Straus MA, Hamby SL, Boney-McCoy S, Sugarman DB. The revised conflict tactics scales (CTS2) development and preliminary psychometric data. Journal of family issues. 1996;17(3):283–316.

31. Koss MP, Abbey A, Campbell R, Cook S, Norris J, Testa M, et al. Revising the SES: A collaborative process to improve assessment of sexual aggression and victimization. Psychology of Women Quarterly. 2007;31(4):357–70.

32. Ahrens KA, Thoma ME, Copen CE, Frederiksen BN, Decker EJ, Moskosky S. Unintended pregnancy and interpregnancy interval by maternal age, National Survey of Family Growth. Contraception. 2018;98(1):52–5.

33. Andrade C. Intent-to-Treat (ITT) vs Completer or Per-Protocol Analysis in Randomized Controlled Trials. Indian Journal of Psychological Medicine. 2022;44(4):416–8.

34. Austin PC, Stuart EA. Moving towards best practice when using inverse probability of treatment weighting (IPTW) using the propensity score to estimate causal treatment effects in observational studies. Statistics in medicine. 2015;34(28):3661–79.

35. Greifer N, Stuart EA, editors. Choosing the Estimand When Matching or Weighting in Observational Studies2021.

36. Nyblade L, Addo NA, Atuahene K, Alsoufi N, Gyamera E, Jacinthe S, et al. Results from a difference-in-differences evaluation of health facility HIV and key population stigma-reduction interventions in Ghana. J Int AIDS Soc. 2020;23(4):e25483.

37. Godard-Sebillotte C, Karunananthan S, Vedel I. Difference-in-differences analysis and the propensity score to estimate the impact of non-randomized primary care interventions. Fam Pract. 2019;36(2):247–51.

38. Uysal J, Schwarz, E., Liambila, W., Wendoh, S., Fonseka, R., Monroy, R.V., Boyce, S.C., Pearson, E., Silverman, J.G. Strategies to address reproductive coercion and intimate partner violence in Nairobi family planning services: qualitative client and provider perspectives. Sexual and Reproductive Health Matters. 2025; In press.

39. Oduenyi C, Obafemi, S., Betron, M. et al. Integrating gender-based violence first-line response into family planning and antenatal care services: an assessment of feasibility, appropriateness and acceptability to healthcare providers in Sokoto and Ebonyi States, Nigeria. BMC Health Services Research. 2025;25:1500.

